# Optimizing Latent Tuberculosis Treatment Strategies Among Immigrants From High-Burden Settings

**DOI:** 10.64898/2026.07.13.26357974

**Authors:** Alexa Tabackman, Meagan Karoly, Karen R. Jacobson, C. Robert Horsburgh, Benjamin Linas, Jeffrey Campbell, Carlos Acuña-Villaorduña, Pranay Sinha

## Abstract

**Importance:** Tuberculosis preventive therapy is central to reducing tuberculosis, and foreign-born individuals account for most US tuberculosis cases. Current US Preventive Services Task Force guidance recommends testing and treating all foreign-born individuals regardless of age or time since immigration, yet the risks of disease progression and of treatment-related harm are not uniform across these groups.

**Objective:** To evaluate the cost-effectiveness and health outcomes of tuberculosis infection treatment strategies among immigrants from high-burden settings, stratified by age and time since immigration.

**Design:** Decision analytical model using individual-level microsimulation (Markov model) over a 30-year horizon, with deterministic and probabilistic (second-order Monte Carlo) sensitivity analyses. Costs and outcomes were discounted at 3%.

**Setting:** US TB and primary care clinics (healthcare-sector perspective), using observed data from the Boston Medical Center/Boston Public Health Commission tuberculosis clinic and published literature.

**Participants:** A simulated cohort of 10 000 IGRA-positive, foreign-born adults from high tuberculosis incidence settings (excluding immunosuppressed individuals), modeled as recent or non-recent (immigrated 25 years earlier) immigrants at ages 35 and 65 years.

**Interventions:** Rifampin daily for 4 months, isoniazid daily for 9 months, or no preventive therapy.

**Main Outcomes and Measures:** Costs, disability-adjusted life-years (DALYs), incident tuberculosis cases and deaths, treatment completion, and incremental cost-effectiveness ratios (ICERs), with the proportion of simulations in which each strategy was optimal at a willingness-to-pay threshold of $50 000 per DALY averted.

**Results:** Among recent immigrants, rifampin was the dominant strategy at ages 35 and 65 years (optimal in 88.5% and 93.9% of simulations), yielding the fewest tuberculosis cases (119.44 and 82.31 per 10 000) and the highest treatment completion (71.4% and 67.7%). Among non-recent immigrants, rifampin remained the dominant strategy (optimal in 53.41% of simulations), followed by no treatment. In 65-year-olds who did not immigrate recently, no treatment was optimal in 94.7% of simulations. ICERs for treatment versus no treatment were unfavorable ($193 600 and $412 857 per DALY averted for rifampin and isoniazid, respectively, at age 65).

**Conclusions and Relevance:** In this decision analytical model, rifampin was cost-effective for recent immigrants, whereas no treatment was optimal for older immigrants with remote arrival date. Age and time since immigration may help risk-stratify tuberculosis infection treatment and reduce unnecessary treatment in lower-risk populations.

**Key Points:** *Question:* Among immigrants from high–tuberculosis-burden settings with a positive interferon-gamma release assay (IGRA), how do rifampin, isoniazid, and no preventive therapy compare in cost effectiveness across age and time since immigration?

*Findings:* In this decision analytical model of 10 000 simulated immigrants, rifampin was the optimal strategy for recent immigrants at ages 35 and 65 years (optimal in 88.5% and 93.4% of simulations), whereas no treatment was optimal for older immigrants who had immigrated 25 years earlier (94.7% at age 65).

*Meaning:* Age and time since immigration may inform tuberculosis infection treatment decisions and reduce unnecessary treatment in lower-risk immigrants.

## Introduction

The global burden of tuberculosis infection is approximately 25% of the world’s population. Addressing TB infection (TBI) with tuberculosis preventive therapy (TPT) is a key component of reducing the incidence of TB in the United States and worldwide.^1–3^ An estimated 13 million people have TB infection in the United States and, without TPT, 1 of 10 will develop TB disease, mainly among foreign born individuals, who account for 70% of all people with TB disease, making diagnosis and treatment of TB infection a crucial public health priority.^4,5^

Despite the availability of effective TPT regimens, completion rates are suboptimal, and there is ongoing debate on which groups should be prioritized for preventive therapy.^6,7^ Current USPSTF guidelines recommend screening with interferon-gamma release assays (IGRA) and offering TPT to all foreign-born individuals regardless of age or time since immigration.^8^ However, the risk of developing TB disease is not homogeneous throughout the lifetime, as rates of progression decline significantly after the first 2 to 5 years post-infection .^9–11^ Furthermore, the low predictive performance of IGRAs for incident TB disease translates into high numbers needed to treat and unnecessary treatments in populations with overall low risk of progression.^12^ TPT also has side effects, especially those related to isoniazid, which increases significantly in individuals older than 65 years.^13^ In this context, the benefit of TPT to prevent progression to TB disease could be outweighed by the low risk of TB reactivation and the higher incidence of side effects, especially in older populations many years after immigration who may undergo routine screening given immigration history.

We conducted a microsimulation modeling study to evaluate the cost-effectiveness and health outcomes of TB infection treatment regimens among immigrants in the United States to better inform clinical decision-making.

## Methods

### Study Design and Study Population

We used observed data from the Boston Medical Center/Boston Public Health Commission (BMC/BPHC) TB clinic to develop a microsimulation model to determine the costs and outcomes of TB infection treatment in a cohort of immigrants from high TB incidence settings with a positive IGRA. The model simulates progression to TB disease in adults over a 30-year time horizon (Table 1). The analysis was conducted from a healthcare-sector perspective, in which TB infection treatment in the United States is provided by TB clinics and primary care. The model excludes patients with moderate-high risk of disease progression, including those with household exposure, HIV infection, receiving tumor necrosis factor inhibitors, chemotherapy, or transplantation and those with diabetes or end-stage renal disease.

**Table 1.** Key Model Assumptions and Input Parameters.

| Parameter | Value (uncertainty interval) | Distribution | Source |
| --- | --- | --- | --- |
| **Costs, \$** | | | |
| Isoniazid (monthly) | 60 (41-80) | Gamma | CDC |
| Rifampin (monthly) | 99 (70-137) | Gamma | CDC |
| TB treatment (monthly) | 3648 (2622-4818) | Gamma | CDC |
| Liver function test | 40 (28-54) | Gamma | BMC/BPHC TB clinic |
| Office visit | 115 (85-155) | Gamma | CMS |
| One hospital day | 1491 (1086-2016) | Gamma | CMS |
| Daily inpatient physician fees | 144 (101-189) | Gamma | CMS |
| **Utilities (disability weights)** |  |  |  |
| Acute TB | 0.33 (0.22-0.45) | PERT | GBD 2019 |
| Drug-induced liver injury (annual) | 0.05 (0.03-0.07) | PERT | GBD 2019 |
| **Probabilities and rates** |  |  |  |
| Rifampin discontinuation, month 1 | 0.11 (0.08-0.14) | Beta | BMC/BPHC clinic |
| Rifampin discontinuation, month 2 | 0.07 (0.04-0.11) | Beta | BMC/BPHC clinic |
| Rifampin discontinuation, month 3 | 0.12 (0.08-0.17) | Beta | BMC/BPHC clinic |
| Isoniazid discontinuation, month 1 | 0.07 (0.05-0.09) | Beta | BMC/BPHC clinic |
| Isoniazid discontinuation, month 2 | 0.05 (0.04-0.08) | Beta | BMC/BPHC clinic |
| Isoniazid discontinuation, month 3 | 0.06 (0.05-0.09) | Beta | BMC/BPHC clinic |
| Isoniazid discontinuation, month 4 | 0.05 (0.02-0.06) | Beta | BMC/BPHC clinic |
| Isoniazid discontinuation, month 5 | 0.06 (0.04-0.10) | Beta | BMC/BPHC clinic |
| Isoniazid discontinuation, month 6 | 0.04 (0.02-0.06) | Beta | BMC/BPHC clinic |
| Isoniazid discontinuation, month 7 | 0.11 (0.08-0.13) | Beta | BMC/BPHC clinic |
| Isoniazid discontinuation, month 8 | 0.08 (0.04-0.11) | Beta | BMC/BPHC clinic |
| Post-treatment TB rate per 100 000 | 2.4 (2.0-2.7) | PERT | Global TB Report 2022 |
| Probability of TB mortality | 0.07 (0.06-0.08) | Beta | Global TB Report 2022 |
| Risk ratio of hepatotoxicity with rifampin | 0.15 (0.069-0.29) | PERT | Menzies et al, 2018 |

Abbreviations: BMC/BPHC, Boston Medical Center/Boston Public Health Commission; CMS, Centers for Medicare & Medicaid Services; GBD, Global Burden of Disease; TB, tuberculosis.

### Model Structure

The Markov model was designed to simulate the natural course of TB infection. The model was constructed as a set of mutually exclusive health states, with natural history transitions represented by movements between states: alive with TB infection, alive with TB disease, death from TB disease, and death from other causes. We used the model to simulate the progression of a cohort of 10 000 IGRA positive patients through these health states in monthly steps over a 30-year projection.

### Model Data

The model assumes that TB exposure occurred immediately prior to immigration and that no further TB exposure occurs in the United States (Table 2). The model incorporated the following treatment strategies: (1) rifampin daily for 4 months (4R) and (2) isoniazid daily for 9 months (9H) for four groups: (1) new 35-year-old immigrants, (2) new 65-year-old immigrants, (3) 35-year-old immigrants who immigrated 25 years previously, and (4) 65-year-old immigrants who immigrated 25 years previously. All strategies were compared with a “do nothing” scenario in which no preventive therapy is offered. Patients could fully or partially complete TB infection treatment. We chose 35 and 65 years because these represent the median age of patients seen at our TB clinic and the age at which liver toxicity increases significantly. Patients could progress to incident TB disease and subsequently complete TB treatment, die from TB, or die from other causes. Probabilities of developing active TB after partial or complete TPT were derived from prior published reports.^14–16^ The risk of progression to active TB disease was modeled as a continuous function of time since immigration, derived from the time-since-infection–specific progression estimates of Menzies et al under the assumption that infection occurred immediately prior to immigration (Table 2). Progression risk was highest shortly after infection and declined monotonically thereafter. The continuous hazard was constructed to reproduce the published cumulative-risk trajectory (3.8%, 6.6%, 7.2%, and 7.9% at 1, 5, 10, and 25 years); the complete single–time-step function used in the model is provided in the Supplement (eTable 3 and eFigure 3).^5,11,17^

**Table 2.** Published Estimates of TB Progression by Time Since Infection (Menzies et al)

| Years since infection | Annual progression rate per 1000 (95% UI) | Cumulative risk of TB, % (95% UI) |
| --- | --- | --- |
| 1 | 38 (33–46) | 3.8 (3.2–4.5) |
| 5 | 3.4 (2.6–4.0) | 6.6 (5.8–7.7) |
| 10 | 0.76 (0.60–0.92) | 7.2 (6.4–8.3) |
| 25 | 0.38 (0.32–0.45) | 7.9 (7.0–8.9) |

Values are annual progression rates per 1000 population, shown as the base-case estimate (low–high bounds). These are representative points from a continuous function of time since immigration, sampled at the midpoints of each interval (2.5, 15, and 27.5 years); the complete function is provided in the Supplement (eTable 3). The model assumes infection occurred immediately prior to immigration, so time since immigration is used as a proxy for time since infection.

Rates of TPT discontinuation and side effects were modeled based on observed data from the BMC/BPHC TB clinic and prior published reports^18^. Age-specific probabilities of adverse events and of hospitalization due to drug-induced liver injury for both the 9H and 4R regimens were modeled as smooth continuous functions of age using quadratic least-squares fits to age-specific estimates obtained from the TSTin3D online interpreter (version 4.0 for adverse events, version 3.0 [2022] for hospitalization), which implements the population-based estimates of Smith et al.^13^ Isoniazid adverse-event and hospitalization risks increase substantially with age, consistent with the well-established age dependence of isoniazid hepatotoxicity; rifampin adverse-event risk increases more modestly with age, while rifampin hospitalization risk remains relatively flat across age groups.^15,19,20^ Full coefficients, data provenance (including interpreter version notes), single-year age lookup tables from 18 to 85 years, and fit diagnostics are provided in the Supplement (eTables 1–2 and eFigures 1–2).

**Figure 1.**
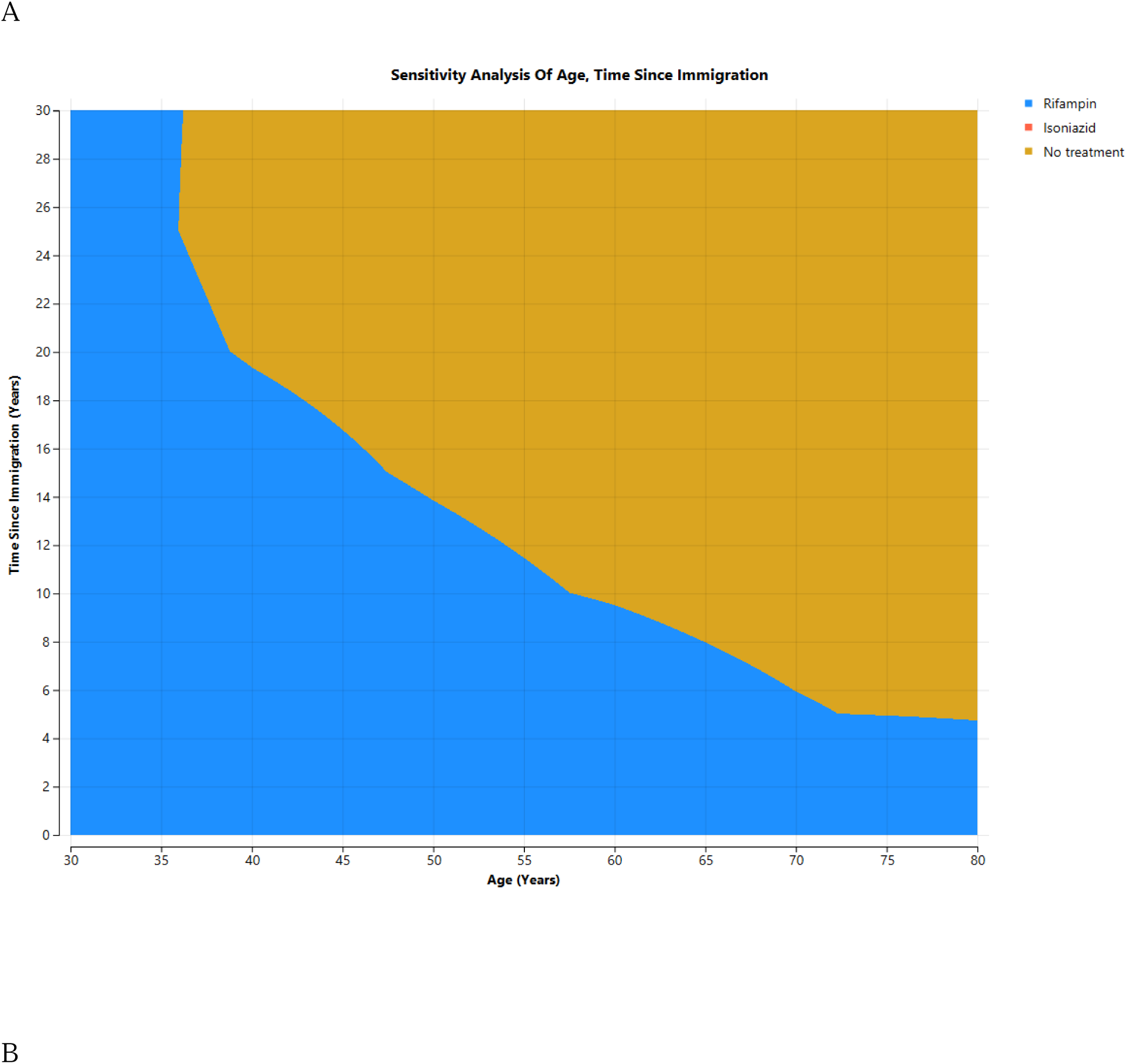
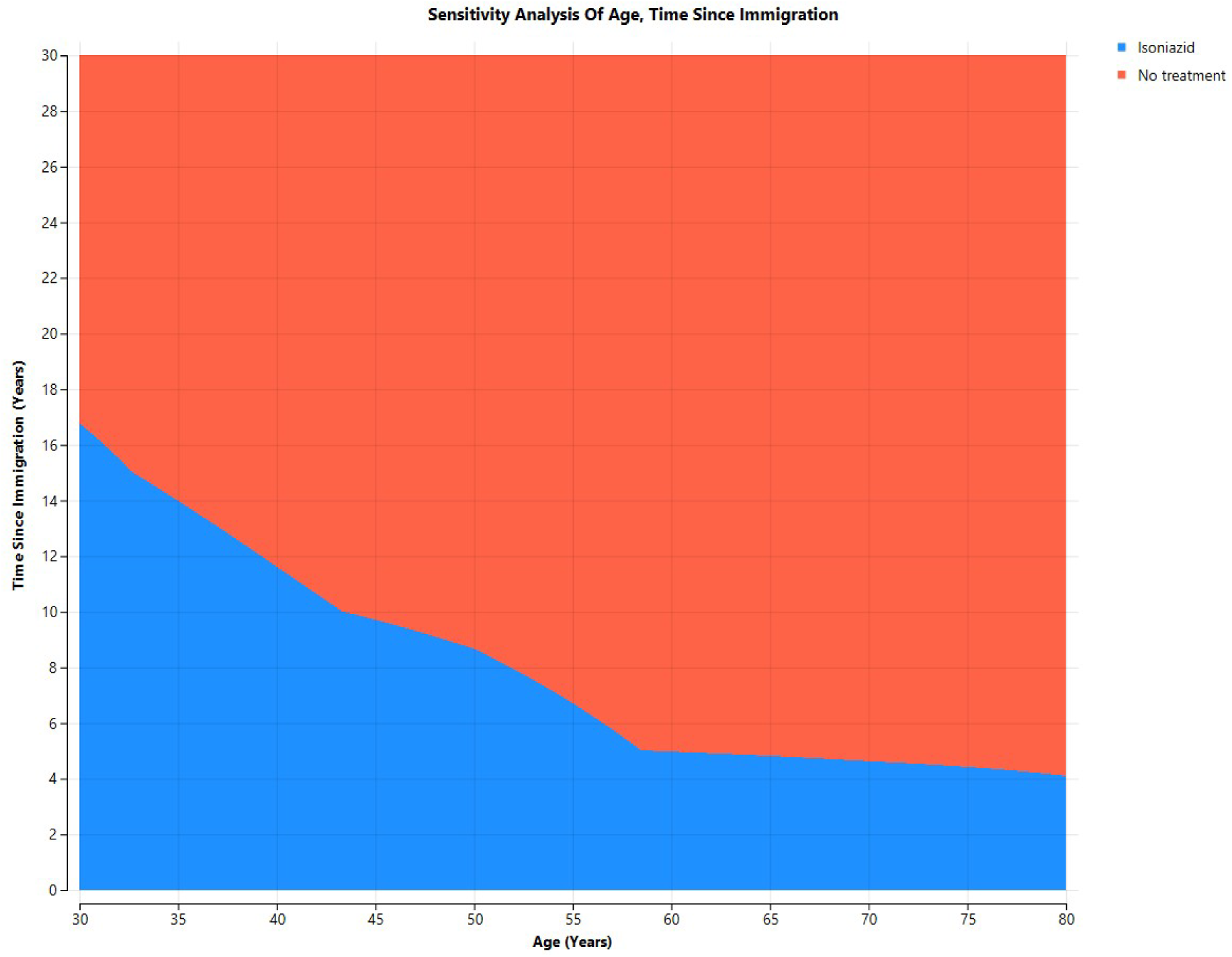
Two-way sensitivity analysis of age at treatment and time since immigration. A, Analysis including all three strategies (rifampin, isoniazid, and no treatment). B, Analysis with rifampin excluded (isoniazid vs no treatment).

**Figure 2.**
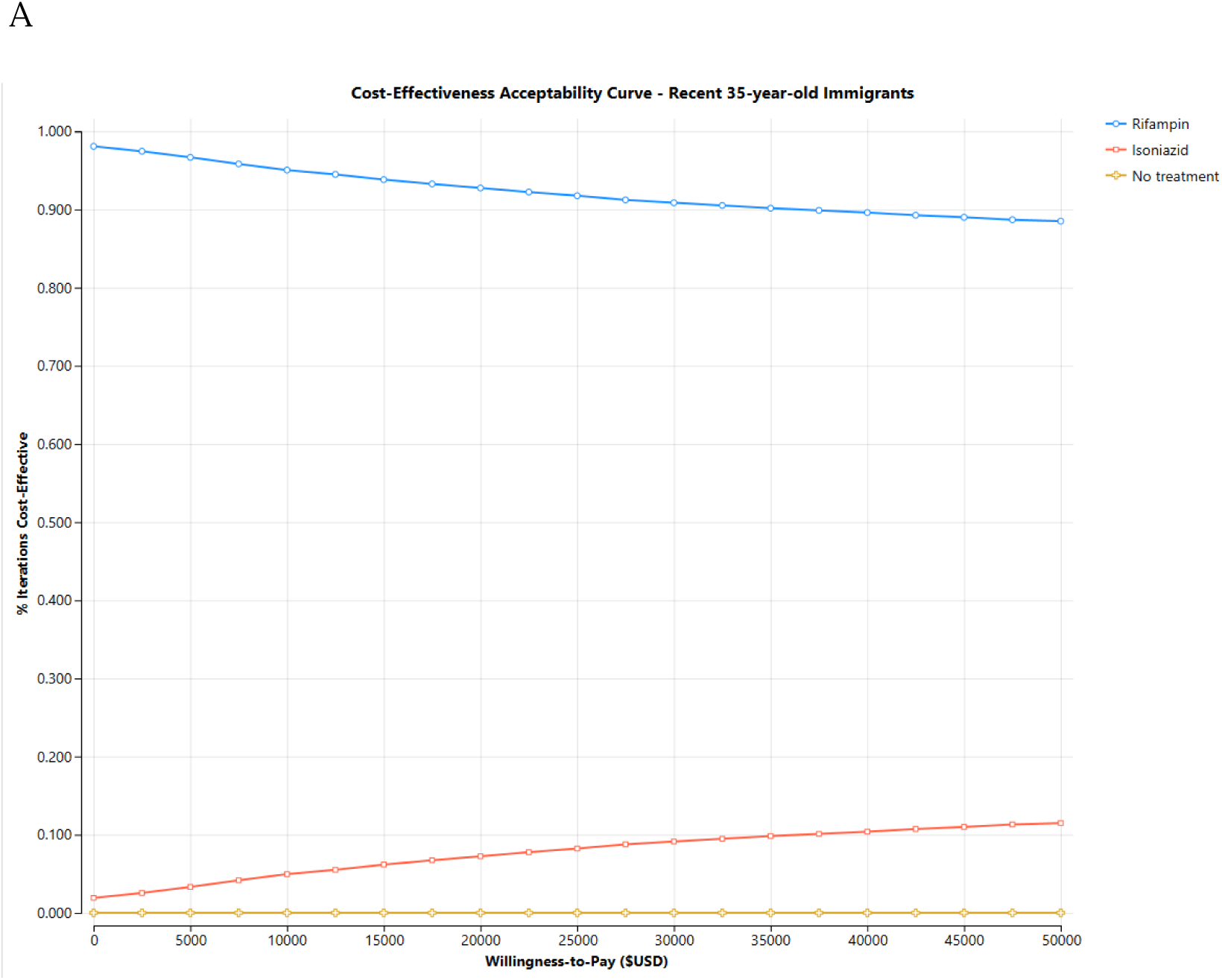
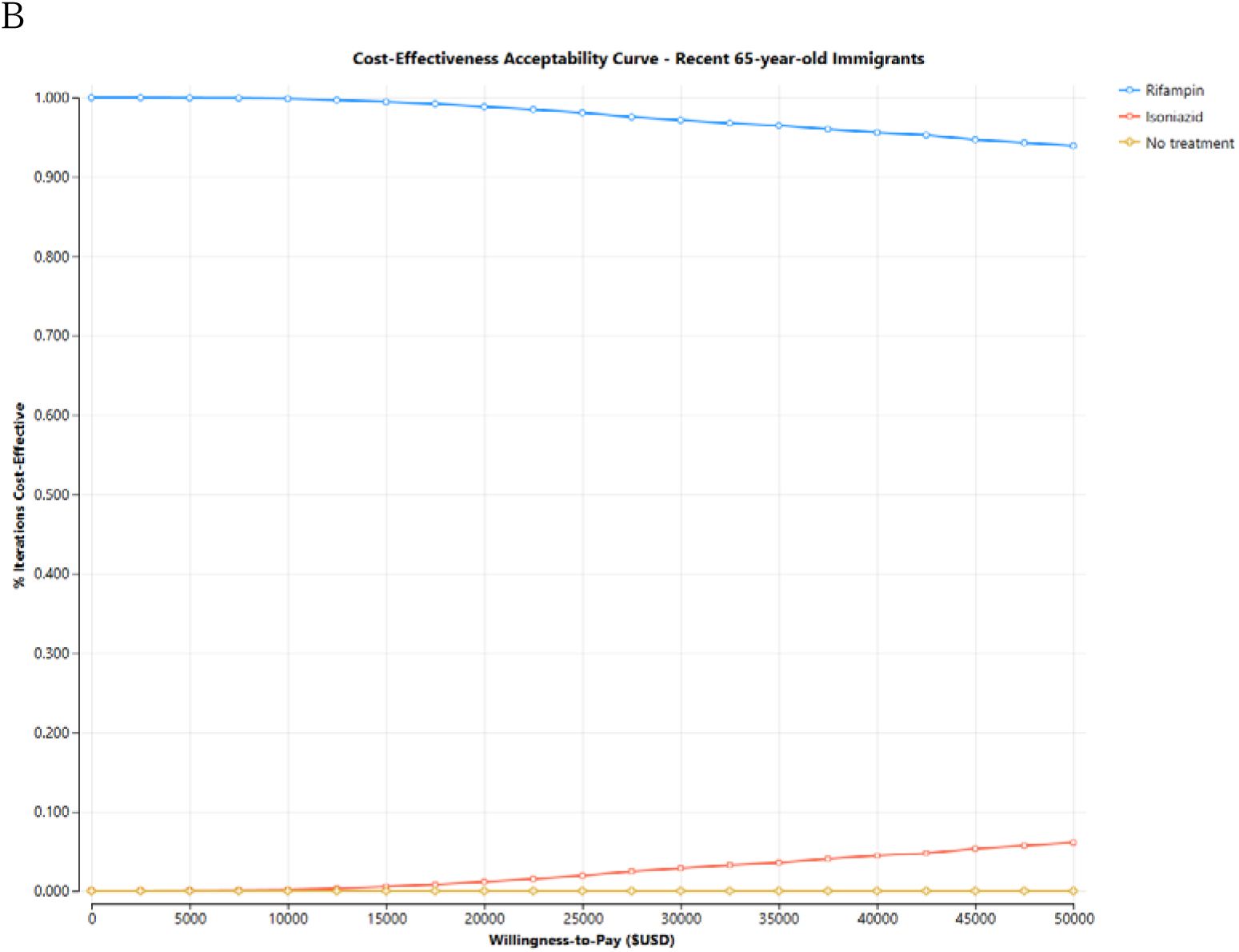
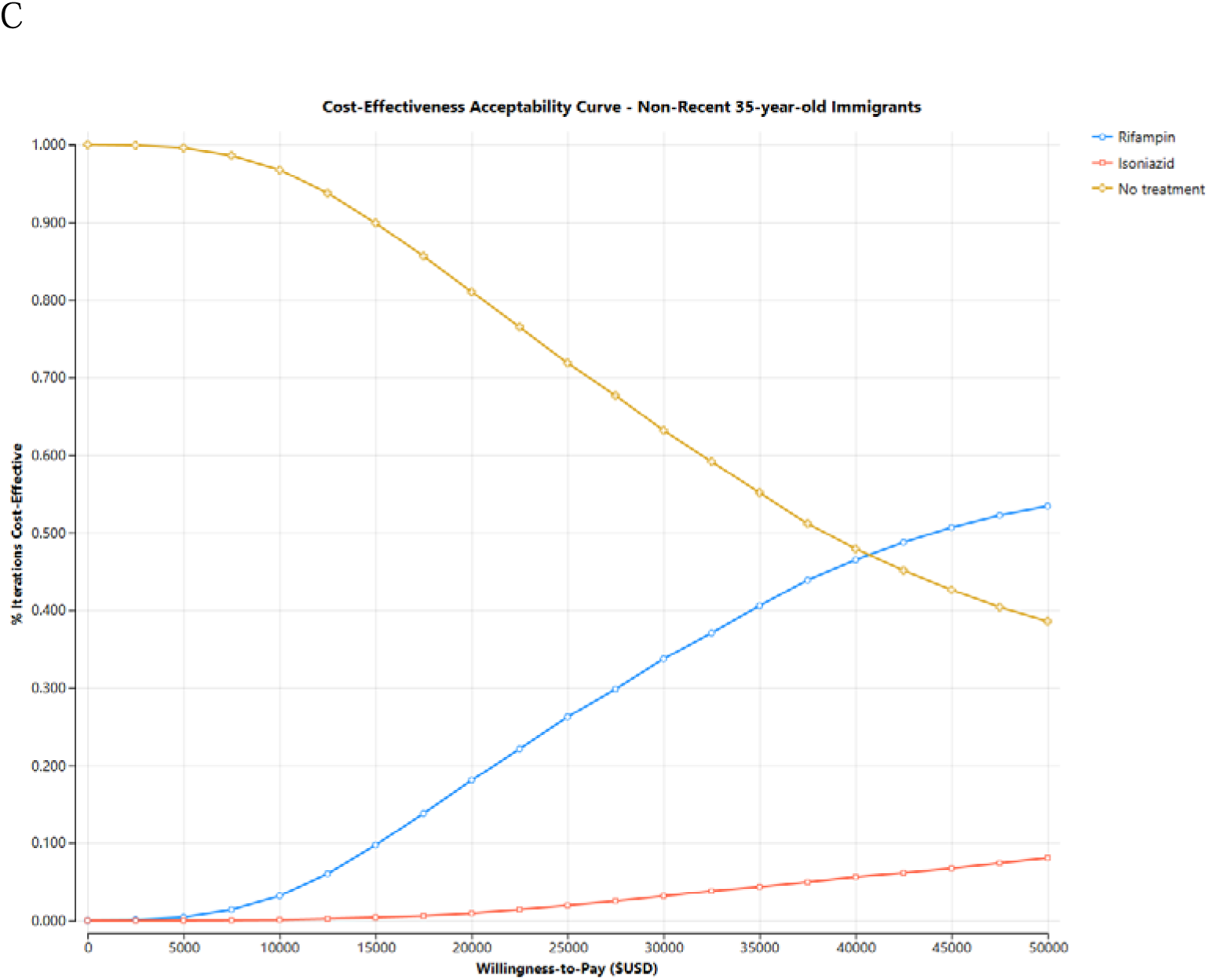
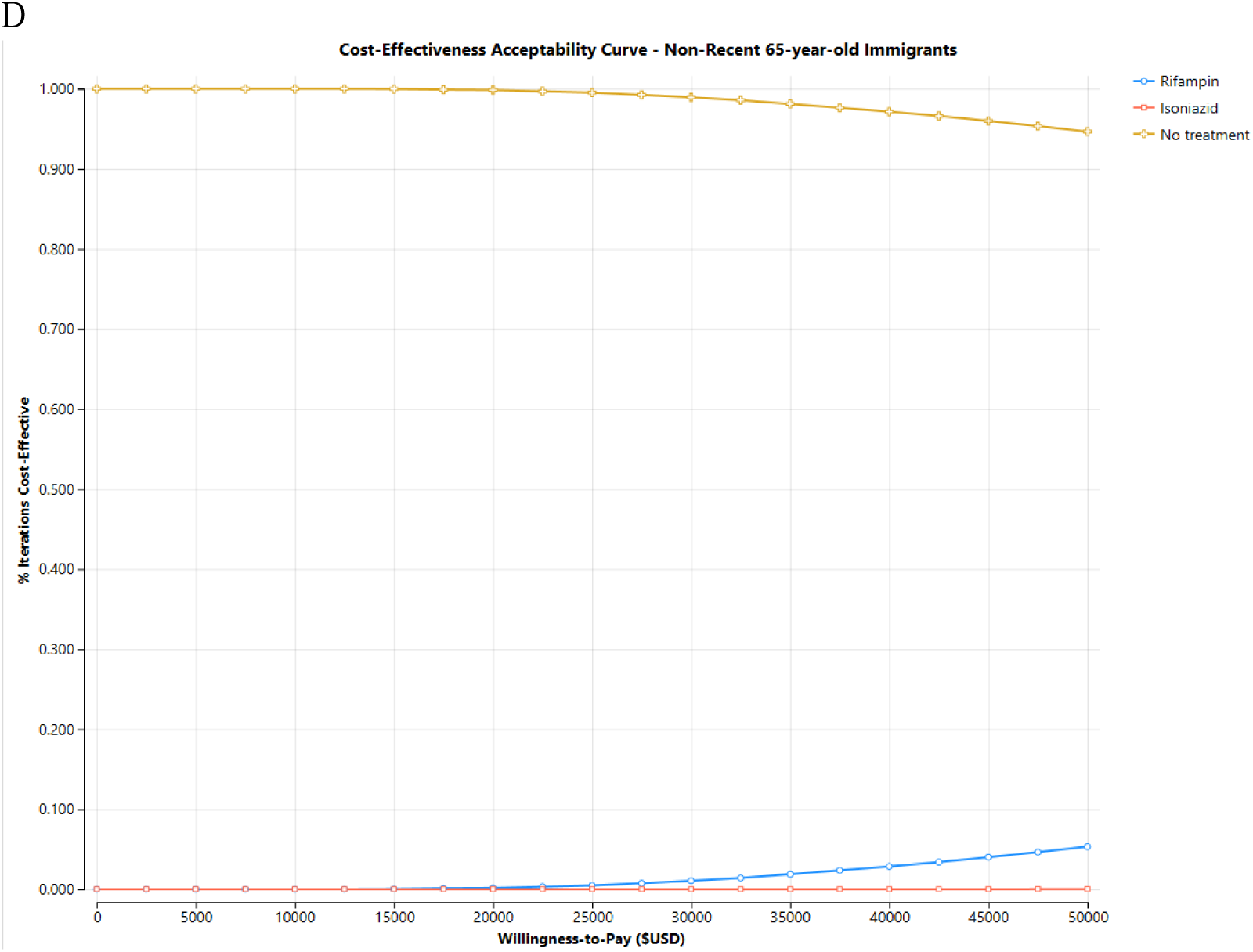
Cost-effectiveness acceptability curves. A–D, Probability that each strategy is cost-effective across willingness-to-pay thresholds for [A] recent 35-year-old, [B] recent 65-year-old, [C] non-recent 35year-old, and [D] non-recent 65-year-old immigrants.

### Simulation

We conducted microsimulation modeling with 10 000 hypothetical IGRA-positive, foreign-born patients. This approach enabled individual-level tracking of outcomes and variability in transition probabilities.

Transition probabilities, treatment completion rates, and adverse event risks were derived from the current literature. Costs were derived from CDC estimates of current TB infection treatment costs.^4^ We simulated hepatotoxicity due to isoniazid and rifampin using published estimates.^20,21^ We additionally incorporated costs associated with clinic visits or hospitalization resulting from adverse effects of isoniazid or rifampin toxicity. We conducted the analysis for each of the four patient subgroups. Costs and outcomes were discounted at 3%.

### Outcomes

Outcomes of interest included cost associated with treatment, rate of progression to active TB, TB deaths, disability-adjusted life-years (DALYs), and percentage completing TB infection treatment.

### Cost-Effectiveness Analysis

We calculated incremental cost-effectiveness ratios (ICERs) as the ratio of mean incremental costs to mean incremental health benefits (DALYs averted) for rifampin and isoniazid treatment compared with no treatment. We used a willingness-to-pay threshold of $50 000.

### Sensitivity Analysis

To assess the robustness of our findings and account for parameter uncertainty, we conducted deterministic and probabilistic sensitivity analyses using second-order Monte Carlo simulation. For each of the four subgroups (recent and non-recent immigrants at ages 35 and 65 years), we ran 10 000 iterations in which key model inputs were varied simultaneously based on probability distributions derived from the published literature (Table 1). We then calculated the proportion of simulations in which each strategy was optimal at a willingness-to-pay threshold of $50 000 per DALY averted. All modeling and analysis were performed using TreeAge Healthcare Pro 2026 (TreeAge Software, Plymouth, MA).

## Results

The outcomes of the simulation model are summarized in Table 3, which presents costs, DALYs, TB cases and deaths, treatment completion rates, and the proportion of simulations in which each strategy was optimal. The cost-effectiveness of the three TB infection treatment strategies by age at treatment and time since immigration is displayed in Figure 1A.

**Table 3.** Model Outcomes by Strategy, Age, and Time Since Immigration.

| Strategy | Mean cost, \$ | DALYs | TB cases per 10 000 | TB deaths per 10 000 | Treatment completion, % | Optimal in simulations, % |
| --- | --- | --- | --- | --- | --- | --- |
| **Recent 35-year-old immigrants** |  |  |  |  |  |  |
| Rifampin | 555.50 | 0.0231 | 119.44 | 8.68 | 71.4% | 88.51% |
| Isoniazid | 739.50 | 0.0293 | 150.74 | 10.88 | 56.5% | 11.49% |
| No treatment | 1462.40 | 0.1332 | 688.80 | 50.10 | N/A | 0.00% |
| **Recent 65-year-old immigrants** |  |  |  |  |  |  |
| Rifampin | 476.70 | 0.0154 | 82.31 | 5.75 | 67.7% | 93.89% |
| Isoniazid | 801.40 | 0.0189 | 100.89 | 6.89 | 50.0% | 6.11% |
| No treatment | 1029.40 | 0.0938 | 502.31 | 34.96 | N/A | 0.00% |
| **Non-recent 35-year-old immigrants** |  |  |  |  |  |  |
| Rifampin | 324.60 | 0.0021 | 10.74 | 0.74 | 71.4% | 53.41% |
| Isoniazid | 447.60 | 0.0027 | 13.38 | 0.96 | 56.5% | 8.05% |
| No treatment | 92.00 | 0.0084 | 43.52 | 3.08 | N/A | 38.54% |
| **Non-recent 65-year-old immigrants** |  |  |  |  |  |  |
| Rifampin | 313.60 | 0.0006 | 2.6 | 0.18 | 67.7% | 5.32% |
| Isoniazid | 601.20 | 0.0007 | 3.14 | 0.23 | 50.0% | 0.01% |
| No treatment | 23.20 | 0.0021 | 11.44 | 0.77 | N/A | 94.67% |

Abbreviations: DALY, disability-adjusted life-year; NA, not applicable; TB, tuberculosis. Remote immigrants immigrated 25 years before the modeled treatment decision. Strategy was considered optimal at a willingness-to-pay threshold of \$50 000 per DALY averted.

Among recent 35-year-old immigrants, rifampin was the dominant strategy compared with isoniazid and no treatment. It resulted in the lowest number of TB cases (119.44 per 10 000), TB deaths (8.68 per 10 000), and DALYs (0.0231), while maintaining the highest treatment completion rate (71.4%). In contrast, isoniazid was associated with higher costs, increased TB cases and deaths, and a lower completion rate (56.5%). The no-treatment strategy led to the highest number of TB cases (688.80), TB deaths (50.10), and DALYs (0.1332). Rifampin was the optimal strategy in 88.5% of simulations.

Among recent 65-year-old immigrants, rifampin again was the preferred strategy, in 93.9% of simulations. It was associated with fewer TB cases (82.31) and deaths (5.75) compared with both isoniazid and no treatment, with a completion rate of 67.7%. While isoniazid also reduced the number of new TB cases, it had a higher cost and lower treatment completion (50.0%) than rifampin. Importantly, isoniazid was the dominant strategy compared with no treatment.

In 35-year-old immigrants who immigrated 25 years previously, rifampin was the optimal strategy in 53.4% of scenarios, followed by no treatment (38.5%). Both isoniazid and rifampin were associated with higher treatment costs than the no-treatment strategy. The costs per DALY averted compared with no treatment were $36 920 for rifampin and $62 385 for isoniazid.

For 65-year-old immigrants who immigrated 25 years previously, no treatment was the optimal strategy in 94.7% of scenarios. Rifampin and isoniazid were both associated with higher treatment costs. The costs per DALY averted were $193 600 and $412 857 for rifampin and isoniazid, respectively, compared to the no treatment strategy.

We additionally conducted a two-way sensitivity analysis of age at treatment and time since immigration in which rifampin was removed as an option. In a comparison of isoniazid with no treatment, no treatment was the optimal strategy for immigrants older than 65 years who had immigrated at least 5 years previously (Figure 1B).

## Discussion

Current guidelines from the CDC and USPSTF recommend screening for TB infection in foreign-born individuals from high-incidence regions and offering treatment to all those with positive testing, but they do not take into account the age of individuals or time since immigration. Our model seeks to address this knowledge gap by comparing strategies for TB infection treatment, accounting for cost-effectiveness, quality of life, and the risk of adverse effects related to TPT.

Our model demonstrated a strong preference for rifampin for treatment of TB infection among recent immigrants in both age groups. Rifampin was associated with higher treatment completion rates, a lower rate of progression to TB disease or death from TB, and greater cost-effectiveness compared with isoniazid in these groups. As such, rifampin should preferentially be used in these populations, with isoniazid reserved for those who cannot tolerate rifampin because of adverse effects or drug-drug interactions. Our model also highlights that time since immigration modifies TB infection progression risk and should be used to inform TB infection treatment decisions. Among immigrants who had immigrated 25 years previously, treatment with rifampin remained the optimal strategy in younger immigrants, however in older non-recent immigrants no treatment was favored, given the higher risk of adverse effects and lower rate of progression to TB disease or death from TB in this population. Our results are similar to those of a recent cost-effectiveness analysis in the Medicare population, in which TPT had a cost per quality-adjusted life-year gained of $112 000 among foreign-born individuals.^22^

Our analysis affirmed that the rate of TB disease and deaths from TB is highest in recent immigrants in both age groups, indicating that testing and treatment for TB infection should be prioritized in this population. In contrast, the low rate of progression to TB disease and TB deaths in older non-recent immigrants indicates that testing and treatment need not be prioritized in this population. Although it is reported that most TB cases in the United States are due to reactivation of latent infection, our model predicts an overall low risk of TB reactivation in non-recent senior immigrants, which is consistent with prior models demonstrating decreased risk of development of TB disease in individuals initially infected many years previously.^11,23^ While current TB incidence rates among non-U.S born individuals in the U.S are highest in those >65, this may reflect those who are more recent arrivals and those at increased risk of disease progression due to immunocompromising conditions.^24^

There are limitations to this analysis. We modeled foreign-born individuals from high-incidence settings under the assumption that no further TB exposure occurred once in the United States, which may underestimate TB risk for certain populations. Nevertheless, TB incidence in most US cities is below 5 per 100 000 cases per year. Notably, we did not include those with significantly increased risk of TB progression, such as those with household exposures, patients with HIV, those receiving immunosuppressive medications, or groups at moderate risk of progression, such as people with diabetes or end-stage renal disease, and these treatment findings cannot be applied to those populations. In addition, we did not include other CDC-recommended shorter TPT regimens, such as weekly isoniazid and rifapentine or daily isoniazid and rifampin for 12 weeks as we did not have data on these regimens from the BMC/BPHC TB clinic. Nevertheless, despite the potential better outcomes of the shorter regimens, their implementation has been slow, and rifampin and isoniazid remain the most commonly used TPT regimens. Finally, progression to TB disease was modeled as a continuous hazard reconstructed from the time-since-infection–specific estimates of Menzies et al and reproducing their published cumulative-risk trajectory; this assumes infection occurred immediately prior to immigration and that time since immigration can be used as time since infection, which may not hold for all immigrants; additionally this does not take into account age at time of infection with regard to risk of progression.^6,25^

Overall, our findings suggest that clinicians who care for immigrant populations may be able to risk stratify their patients with TB infection based on age and time since immigration to determine who will best benefit from treatment. This approach has the potential to reduce unnecessary treatment in low-risk populations and optimize health outcomes. Additionally, this model can provide a basis for future observational studies to further address this knowledge gap and inform updated recommendations for TB infection treatment in the United States.

## Data Availability

All data produced in the present work are contained within the manuscript and supplementary appendix

## Article Information

### Data Sharing Statement

All relevant data is reported within the manuscript and Supplementary Appendix

### Ethics/IRB

This analysis was exempt from IRB approval at Boston Medical Center

### Reporting Guideline

This study follows the CHEERS reporting guideline for economic evaluations

