## Supplemental Materials for "Optimizing Latent Tuberculosis Treatment Strategies Among Immigrants From High-Burden Settings"

**Supplementary Appendix: Smoothed age-dependent probabilities of adverse events and hospitalization for 9H and 4R latent tuberculosis preventive therapy**

### **S1. Rationale**

In the base model, age-specific probabilities of adverse events and of hospitalization for isoniazid (9H) and rifampin (4R) were specified as step functions across discrete age strata. This produces implausible discontinuities at the boundaries of those strata, under which two patients differing by a single year of age can be assigned meaningfully different risks. To address this, we re-modeled each age-dependent probability as a smooth continuous function of age, anchored on the same underlying clinic-observed data points used to parameterize the step-function version.

### **S2. Data sources and provenance**

All anchor values were obtained from the TSTin3D online interpreter (https://www.tstin3d.com), a widely used risk-estimation tool developed and maintained by the McGill International TB Centre. The interpreter's adverse-event module implements the population-based estimates of Smith et al. (reference 11 of the main manuscript), which remain the primary peer-reviewed source for the underlying probabilities.

The present model was initially constructed in 2022 using Version 3.0 of the interpreter. Between the original model build and the present analysis, the interpreter was updated to Version 4.0. Two changes between versions are relevant here. First, v4.0 revised the age-specific adverse-event probabilities and, in particular, introduced age-based increases in rifampin adverse-event risk that were not present in v3.0, which treated rifampin toxicity as essentially age-independent. Second, v4.0 removed the hospitalization module entirely and no longer reports age-specific hospitalization probabilities for either regimen.

For the present analysis we therefore adopted a mixed-provenance approach. Age-specific adverse-event probabilities for both 9H and 4R were taken from the updated v4.0 interpreter, because retaining the v3.0 values — under which rifampin adverse-event risk did not increase with age — would have systematically biased the cost-effectiveness comparison in favor of rifampin relative to isoniazid in older patients. Age-specific hospitalization probabilities for INH and RIF were taken from v3.0 (as accessed in 2022), because v4.0 does not provide equivalents. We contacted the tool maintainers to request clarification on the rationale for the v3.0→v4.0 changes and on whether the underlying source data for hospitalization were updated.

The following anchor values were used for the quadratic fits:

Adverse events (v4.0): 9H — 0.030 at age 18, 0.047 at age 35, 0.068 at age 51, 0.063 at age 66; 4R — 0.017 at age 18, 0.021 at age 35, 0.024 at age 51, 0.041 at age 66.

Hospitalization due to drug-induced adverse events (v3.0, 2022): INH — 0.001 at age 35, 0.002 at age 40, 0.006 at age 50, 0.024 at age 65; RIF — 0.00070 at age 35, 0.00050 at age 40, 0.00060 at age 50, 0.00140 at age 65.

### **S3. Functional form and fitting**

For each outcome we fit a quadratic of the form P(age) = a + b·age + c·age² using ordinary nonlinear least squares over the anchor points (scipy.optimize.curve_fit). A quadratic was chosen because it is the lowest-order polynomial capable of capturing the observed non-monotonic behavior of the 9H adverse-event data (which peaks in late middle age before declining slightly) and the U-shaped pattern of the rifampin hospitalization data, while remaining robust to the sparse number of anchor points (four per curve).

Fitted coefficients:

• P(adverse event | 9H): a = -8.1482e-3, b = 2.3892e-3, c = -1.9465e-5

• P(adverse event | 4R): a = 2.5193e-2, b = -6.5400e-4, c = 1.3333e-5

• P(hospitalization | INH): a = 3.8917e-2, b = -2.0621e-3, c = 2.8182e-5

• P(hospitalization | RIF): a = 4.5167e-3, b = -1.8182e-4, c = 2.0606e-6

### **S4. Handling of ages outside the anchor range**

The adverse-event data span ages 18 to 66; the hospitalization data span ages 35 to 65. Because quadratic extrapolation beyond the anchor range can produce implausible or non-monotonic values that reflect curvature artifacts rather than biology, we clamp each fitted function at its value at the nearest anchor-range endpoint. Specifically: for adverse events, the age-18 fit value is used for any age below 18 (not applicable in this model) and the age-66 fit value is used for ages above 66; for hospitalization, the age-35 fit value is used for ages 18–34 and the age-65 fit value is used for ages 66–85. This choice is conservative in the sense that it avoids projecting the 9H adverse-event curve's late-life apparent decline — which is driven by a single anchor point at age 66 — into ages for which no data exist. Sensitivity analyses could alternatively allow free quadratic extrapolation or substitute a monotonic functional form (e.g. logistic in age) if desired.

### **S5. Fit diagnostics**

Figures S1 and S2 show the quadratic fits overlaid on the anchor points. Solid lines indicate the fitted quadratic within the anchor range; dashed lines and shaded regions indicate ages at which values are clamped at the nearest in-range fitted value. The 9H quadratic in Figure S1 does not pass exactly through the age-51 and age-66 anchors because the least-squares fit balances residuals across all four points; the mild non-monotonicity of the raw 9H anchors (age-66 value below age-51 value) is therefore smoothed by the fit, which peaks around age 61 and then declines very slightly. In Figure S2, the steep age gradient of INH hospitalization risk is clearly visible and contrasts with the much flatter RIF hospitalization curve, consistent with the clinical understanding that isoniazid hepatotoxicity is strongly age-dependent while rifampin hospitalization risk is much less so.

**Figure S1. Quadratic fits for age-specific probability of adverse events on 9H and 4R regimens, with TSTin3D v4.0 anchor points. Shaded region (age > 66) indicates clamped extrapolation.**

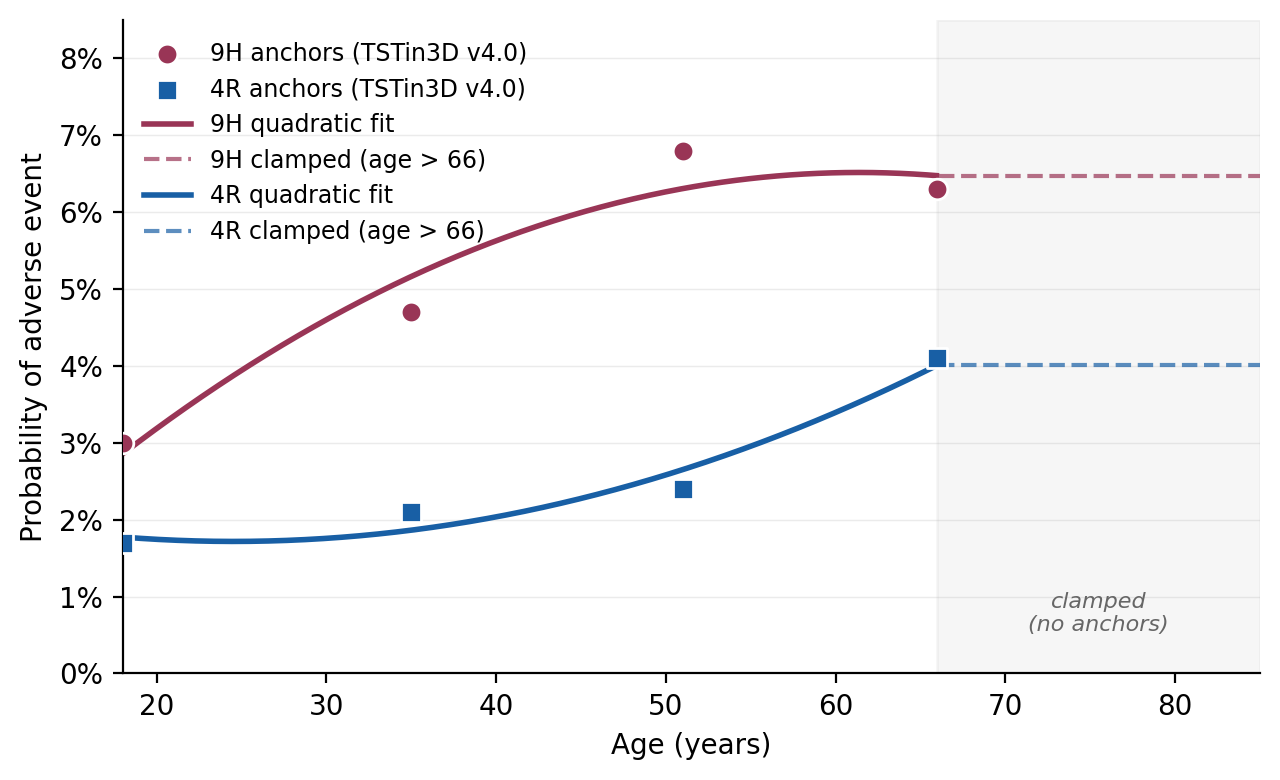

**Figure S2. Quadratic fits for age-specific probability of hospitalization on INH and RIF, with TSTin3D v3.0 (2022) anchor points. Shaded regions (age < 35 and age > 65) indicate clamped extrapolation.**

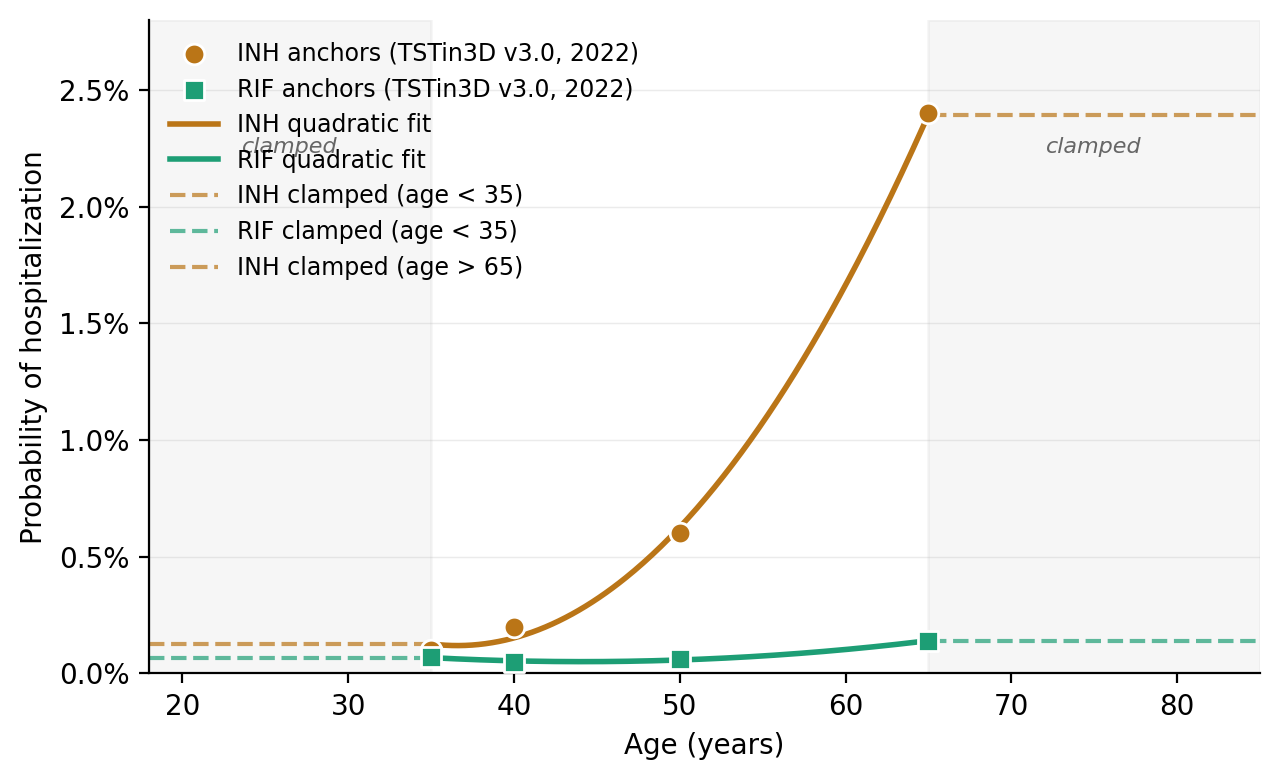

### **S6. Lookup tables**

**Table S1. Age-specific probability of any adverse event while on 9H or 4R latent tuberculosis preventive therapy, smoothed quadratic fit with clamping outside the anchor range (ages 18–66).**

| **Age (years)** | **P(adverse event \| 9H)** | **P(adverse event \| 4R)** |
| --- | --- | --- |
| 18 | 0.0286 | 0.0177 |
| 19 | 0.0302 | 0.0176 |
| 20 | 0.0318 | 0.0174 |
| 21 | 0.0334 | 0.0173 |
| 22 | 0.0350 | 0.0173 |
| 23 | 0.0365 | 0.0172 |
| 24 | 0.0380 | 0.0172 |
| 25 | 0.0394 | 0.0172 |
| 26 | 0.0408 | 0.0172 |
| 27 | 0.0422 | 0.0173 |
| 28 | 0.0435 | 0.0173 |
| 29 | 0.0448 | 0.0174 |
| 30 | 0.0460 | 0.0176 |
| 31 | 0.0472 | 0.0177 |
| 32 | 0.0484 | 0.0179 |
| 33 | 0.0495 | 0.0181 |
| 34 | 0.0506 | 0.0184 |
| 35 | 0.0516 | 0.0186 |
| 36 | 0.0526 | 0.0189 |
| 37 | 0.0536 | 0.0192 |
| 38 | 0.0545 | 0.0196 |
| 39 | 0.0554 | 0.0200 |
| 40 | 0.0563 | 0.0204 |
| 41 | 0.0571 | 0.0208 |
| 42 | 0.0579 | 0.0212 |
| 43 | 0.0586 | 0.0217 |
| 44 | 0.0593 | 0.0222 |
| 45 | 0.0599 | 0.0228 |
| 46 | 0.0606 | 0.0233 |
| 47 | 0.0611 | 0.0239 |
| 48 | 0.0617 | 0.0245 |
| 49 | 0.0622 | 0.0252 |
| 50 | 0.0626 | 0.0258 |
| 51 | 0.0631 | 0.0265 |
| 52 | 0.0635 | 0.0272 |
| 53 | 0.0638 | 0.0280 |
| 54 | 0.0641 | 0.0288 |
| 55 | 0.0644 | 0.0296 |
| 56 | 0.0646 | 0.0304 |
| 57 | 0.0648 | 0.0312 |
| 58 | 0.0649 | 0.0321 |
| 59 | 0.0651 | 0.0330 |
| 60 | 0.0651 | 0.0340 |
| 61 | 0.0652 | 0.0349 |
| 62 | 0.0652 | 0.0359 |
| 63 | 0.0651 | 0.0369 |
| 64 | 0.0650 | 0.0379 |
| 65 | 0.0649 | 0.0390 |
| 66 | 0.0647 | 0.0401 |
| 67 | 0.0647 | 0.0401 |
| 68 | 0.0647 | 0.0401 |
| 69 | 0.0647 | 0.0401 |
| 70 | 0.0647 | 0.0401 |
| 71 | 0.0647 | 0.0401 |
| 72 | 0.0647 | 0.0401 |
| 73 | 0.0647 | 0.0401 |
| 74 | 0.0647 | 0.0401 |
| 75 | 0.0647 | 0.0401 |
| 76 | 0.0647 | 0.0401 |
| 77 | 0.0647 | 0.0401 |
| 78 | 0.0647 | 0.0401 |
| 79 | 0.0647 | 0.0401 |
| 80 | 0.0647 | 0.0401 |
| 81 | 0.0647 | 0.0401 |
| 82 | 0.0647 | 0.0401 |
| 83 | 0.0647 | 0.0401 |
| 84 | 0.0647 | 0.0401 |
| 85 | 0.0647 | 0.0401 |

**Table S2. Age-specific probability of hospitalization attributable to drug-induced adverse events while on INH or RIF, smoothed quadratic fit with clamping outside the anchor range (ages 35–65).**

| **Age (years)** | **P(hospitalization \| INH)** | **P(hospitalization \| RIF)** |
| --- | --- | --- |
| 18 | 0.00127 | 0.00068 |
| 19 | 0.00127 | 0.00068 |
| 20 | 0.00127 | 0.00068 |
| 21 | 0.00127 | 0.00068 |
| 22 | 0.00127 | 0.00068 |
| 23 | 0.00127 | 0.00068 |
| 24 | 0.00127 | 0.00068 |
| 25 | 0.00127 | 0.00068 |
| 26 | 0.00127 | 0.00068 |
| 27 | 0.00127 | 0.00068 |
| 28 | 0.00127 | 0.00068 |
| 29 | 0.00127 | 0.00068 |
| 30 | 0.00127 | 0.00068 |
| 31 | 0.00127 | 0.00068 |
| 32 | 0.00127 | 0.00068 |
| 33 | 0.00127 | 0.00068 |
| 34 | 0.00127 | 0.00068 |
| 35 | 0.00127 | 0.00068 |
| 36 | 0.00120 | 0.00064 |
| 37 | 0.00120 | 0.00061 |
| 38 | 0.00125 | 0.00058 |
| 39 | 0.00136 | 0.00056 |
| 40 | 0.00152 | 0.00054 |
| 41 | 0.00174 | 0.00053 |
| 42 | 0.00202 | 0.00052 |
| 43 | 0.00235 | 0.00051 |
| 44 | 0.00274 | 0.00051 |
| 45 | 0.00319 | 0.00051 |
| 46 | 0.00369 | 0.00051 |
| 47 | 0.00425 | 0.00052 |
| 48 | 0.00487 | 0.00054 |
| 49 | 0.00554 | 0.00056 |
| 50 | 0.00627 | 0.00058 |
| 51 | 0.00705 | 0.00060 |
| 52 | 0.00789 | 0.00063 |
| 53 | 0.00879 | 0.00067 |
| 54 | 0.00974 | 0.00071 |
| 55 | 0.01075 | 0.00075 |
| 56 | 0.01182 | 0.00080 |
| 57 | 0.01294 | 0.00085 |
| 58 | 0.01412 | 0.00090 |
| 59 | 0.01535 | 0.00096 |
| 60 | 0.01664 | 0.00103 |
| 61 | 0.01799 | 0.00109 |
| 62 | 0.01940 | 0.00116 |
| 63 | 0.02086 | 0.00124 |
| 64 | 0.02237 | 0.00132 |
| 65 | 0.02395 | 0.00140 |
| 66 | 0.02395 | 0.00140 |
| 67 | 0.02395 | 0.00140 |
| 68 | 0.02395 | 0.00140 |
| 69 | 0.02395 | 0.00140 |
| 70 | 0.02395 | 0.00140 |
| 71 | 0.02395 | 0.00140 |
| 72 | 0.02395 | 0.00140 |
| 73 | 0.02395 | 0.00140 |
| 74 | 0.02395 | 0.00140 |
| 75 | 0.02395 | 0.00140 |
| 76 | 0.02395 | 0.00140 |
| 77 | 0.02395 | 0.00140 |
| 78 | 0.02395 | 0.00140 |
| 79 | 0.02395 | 0.00140 |
| 80 | 0.02395 | 0.00140 |
| 81 | 0.02395 | 0.00140 |
| 82 | 0.02395 | 0.00140 |
| 83 | 0.02395 | 0.00140 |
| 84 | 0.02395 | 0.00140 |
| 85 | 0.02395 | 0.00140 |

### **S7. Implementation notes and limitations**

These lookup tables are intended to be used directly by the microsimulation: at each simulated patient's current age, the corresponding probability is read from Table S1 (adverse event) or Table S2 (hospitalization given adverse event) and applied as a per-event Bernoulli draw. Because the underlying functions are smooth in age, the model output is now continuous in age at baseline, removing the artifactual jumps at the 40/50/65 strata boundaries that were present in the original parameterization.

Two limitations of this parameterization deserve explicit mention. First, the adverse-event and hospitalization anchors derive from different versions of the TSTin3D interpreter (v4.0 and v3.0 respectively), as described in Section S2. Although both versions ultimately trace to the same peer-reviewed source literature, the exact fitting and interpolation rules used by the interpreter differ between versions and are not fully published. As a sensitivity check, the model can be re-run using v3.0 adverse-event values to bracket the impact of this choice. Second, the TSTin3D interpreter was designed for use in adults under age 80, and both of our anchor data sets end below that age (age 66 for adverse events; age 65 for hospitalization). Values reported in the lookup tables for ages 80–85 are therefore conservative floor estimates obtained by clamping at the upper-anchor fitted value, and may underestimate true adverse-event and hospitalization risk at the oldest ages. Given that older remote immigrants are the subgroup in which our model already finds no-treatment to be optimal, this conservatism is unlikely to qualitatively change the main conclusions, but would tend to further strengthen the case against treatment in that subgroup if corrected.
